# Carotid Plaque Calcification Associates with Cardiac Morbidity, Mortality and Systemic Atherosclerotic Disease in a Sex-Specific Manner

**DOI:** 10.64898/2026.07.07.26357513

**Authors:** Henna Komi, Petra Ijäs, Krista Nuotio, Riikka Tulamo, Henrietta Törmänen, Ella Rytkölä, Laura Mäkitie, Perttu Lindsberg, Lauri Soinne, Marja-Liisa Lokki, Pirkka Vikatmaa, Mohammadreza Shoghli, Mikko I. Mäyränpää, Inkeri Lokki, Juha Sinisalo

## Abstract

**Background:** Carotid plaque calcification is commonly interpreted as a marker of atherosclerotic burden, but its prognostic meaning may depend on calcification morphology. Whether histologically defined calcification subtypes in carotid plaques identify systemic cardiovascular disease and long-term cardiac risk remains unclear.

**Methods:** We studied 479 patients, including 154 women, undergoing carotid endarterectomy in the Helsinki Carotid Endarterectomy Study 2 with 10-year follow-up. Hematoxylin–eosin-stained plaque sections were digitized, and artificial intelligence–based image analysis was used to quantify sheet, nodular, and total calcification as proportions of plaque tissue area. The primary endpoint was cardiac death; secondary endpoints were major coronary events and major adverse limb events (MALE). Associations between calcification tertiles and outcomes were assessed using Fine–Gray regression models adjusted for conventional cardiovascular risk factors, with prespecified sex-stratified analyses.

**Results:** Total calcification was associated with baseline coronary artery disease and chronic heart failure but did not predict cardiac mortality. In contrast, nodular calcification was independently associated with cardiac death. Compared with the lowest tertile, the middle and highest tertiles of nodular calcification were associated with increased cardiac mortality in the whole cohort (subdistribution hazard ratio [sHR], 2.87 [95% CI, 1.39–5.91] and 2.55 [95% CI, 1.24–5.27], respectively). Nodular calcification was also associated with baseline peripheral artery disease. High sheet calcification was associated with cardiac death in men (sHR, 2.3 [95% CI, 1.0–5.1]) but not in women. Exploratory sex-stratified analyses suggested that high total calcification was associated with major coronary events and MALE in women, but not in men.

**Conclusions:** Carotid plaque calcification morphology, rather than total calcification burden alone, is associated with long-term cardiac mortality after carotid endarterectomy. Nodular calcification emerged as the strongest prognostic phenotype, while exploratory sex-stratified findings suggest that total calcification may reflect coronary and peripheral event risk in women.

## Introduction

Carotid atherosclerosis affects more than 800 million individuals aged 30–79 years worldwide. Its prevalence increases markedly with age, such that by 70 years, carotid plaque is present in 60.8% of men and 45.0% of women.^1^ The most important determinant of prognosis in patients with atherosclerosis is the coexistence of coronary artery disease (CAD), which remains the leading cause of death globally, accounting for approximately 9.0 million deaths annually.^2^ Atherosclerosis is a systemic disease typically affecting multiple vascular territories, and more than 60% of patients with carotid atherosclerosis are estimated to have underlying, often asymptomatic CAD.^3–6^ Furthermore, the overlap between carotid atherosclerosis and CAD with lower extremity peripheral artery disease (PAD) is conspicuous.^7,8^

The presence and severity of carotid atherosclerosis are associated with an increased risk of myocardial infarction.^9,10^ Patients with advanced atherosclerosis, including those with peripheral arterial involvement, are at particularly high risk of myocardial infarction and cardiac death.^11–13^ Plaque instability in one vascular bed has been proposed to reflect vulnerability of plaques in other arterial territories. Supporting this concept, carotid plaques from patients presenting with acute coronary syndrome have been shown to exhibit unstable features.^14,15^ These observations suggest that assessment of systemic plaque characteristics, rather than isolated vascular territories, may improve cardiovascular risk stratification.

Carotid plaque calcification has been associated with CAD in a cross-sectional study,^16^ and several studies have reported a suggestive association between carotid plaque calcification and myocardial infarction as well as peripheral arterial events.^17,18^ Carotid calcification has also been linked to hypertension.^19^ However, longitudinal studies incorporating histological calcification quantification including calcification subtype analysis and long-term cardiovascular outcomes are lacking. Mechanistically, arterial calcification is a heterogenous process, and its association with plaque stability appears to depend on calcification morphology. Large, dense calcifications have been suggested to be a feature of stability locally in carotid plaques^20–22^ and coronary plaques^23,24^. Histologically, calcification can be classified into distinct subtypes, primarily sheet calcification and nodular calcification.^25^ Sheet calcification consists of large, homogeneous calcium plates and has been suggested to be more prevalent in stable plaques.^21^ Nodular calcification, characterized by fragmented calcium deposits surrounded by fibrin, is proposed to arise from fractured sheet calcifications but also from resolving thrombi.^23,26^

The onset and progression of atherosclerosis differ between sexes. Women typically develop atherosclerotic disease 10-20 years later than men; however, the disease burden and morbidity increase rapidly after menopause.^27–29^ In addition, sex-related disparities exist in plaque pathophysiology, with male plaques more frequently exhibiting vulnerable features.^30–36^ Despite these differences, most studies on carotid plaque calcification have not performed sex-stratified analyses.

Therefore, we aimed to investigate the prognostic relevance of carotid plaque calcification and its histologically defined subtypes for long-term cardiovascular outcomes in patients undergoing carotid endarterectomy, with a particular focus on identifying potential sex-specific associations.

## Methods

### Study population and sample collection

The Helsinki Carotid Endarterectomy Study (HeCES2) is a single-center, prospective, cohort study including 500 consecutive patients undergoing carotid endarterectomy (CEA) between October 2012 to September 2015 at Helsinki University Hospital, Finland. The study design has been previously published.^37^

Patients were referred for CEA according to guidelines from the European Stroke Organisation and the European Society of Vascular Surgery.^38,39^ The study was approved by the local Ethics Committee (reference number 147/13/03/01/2011) and hospital research board. All participants provided written informed consent.

Patients underwent baseline interviews and clinical examinations, and medical records were reviewed for the history of CAD and/or coronary artery bypass grafting (CABG) and other diagnosed comorbidities including PAD. Carotid plaques obtained during CEA were evaluated macroscopically and histologically. Description of plaque tissue processing has previously been described.^40^ Hematoxylin-eosin (HE) staining of 479 patients’ plaques was accessible for further analysis with artificial intelligence (AI) -driven calcification quantification.

### Aiforia Artificial Intelligence Algorithm

The development, training and validation of the deep learning algorithm to quantify two classes of arterial calcification has previously been described.^40^ Hematoxylin and eosin-stained slides of longitudinal carotid plaque sections were digitized, and the quantification of nodular and sheet calcification was performed by a cloud-based image deep learning algorithm (Aiforia Create, Aiforia Technologies Oy, Helsinki, Finland, https://www.aiforia.com/). The algorithm quantified the areas of nodular calcification and sheet calcification both in mm^2^ and as a proportion of the calcification category to that of the plaque section tissue area (%). Proportional measurement of calcification area from total tissue area was used for downstream analyses. Total calcification area (%) was calculated as the sum of nodular calcification (%) and sheet calcification (%).

### Outcome variables

Follow-up began 30 days after the CEA to minimize confounding from perioperative complications and immediate postoperative events. Data was collected from electronic medical records in the autumn of 2025. If alive at the date of data collection, the patient was censored at his/her last visit to healthcare at the hospital district. The primary endpoint was cardiac death. Secondary endpoints included major coronary events and major adverse limb events (MALE). Major coronary events were defined as myocardial infarction (either ST-elevation myocardial infarction or non-ST-elevation myocardial infarction), or need for revascularization (coronary artery bypass grafting (CABG) or percutaneous coronary angioplasty (PCI) with or without stent placement). Major adverse limb events (MALE) were defined as open or endovascular surgery for symptomatic PAD of lower limb (severely disabling intermittent claudication, chronic limb threatening ischemia, acute limb ischemia) or major amputation due to severe lower limb ischemia.

Death certificates dated before the end of 2022 were obtained from the Finnish public authority Statistics Finland.^41^ Mortality data including the dates from 2023 onwards was gathered from patients’ electronic medical records, which also included a death certificate for patients located in the local hospital district at the time of death. Death was categorized as cardiac death or death by other reasons utilizing diagnoses for death and the descriptive information in the death certificate or patient records. In case of ambiguity, a consensus meeting was held between K.N., P.I., R.T., H.K., and M.I.M to determine the final cause of death.

### Statistical Analysis

Non-normally distributed variables are presented as median (interquartile range), and normally distributed variables are given as mean ± standard deviation (SD). The Mann-Whitney U test was used for non-normally distributed continuous variables, and the t-test for normally distributed continuous variables. For binary variables, the chi-square test was used.

Calcification variables were divided into sex-specific tertiles: T1 [low], T2 [mid], and T3 [high]. Statistical significance was defined as two-sided p < 0.05 in all analyses. All statistical analyses were performed using R version RStudio version 2023.12.1 and SPSS version 29.0.0.0.

### Linear regression models

Associations between calcification measures and clinical covariates were examined using linear regression models. All analyses were performed using one-variable-at-a-time models with age adjustment. For sex-stratified analyses, models were fitted separately in men and women with the calcification outcome as the dependent variable and a single covariate as the independent variable. For overall analyses, sex was included as an additional covariate in the model. Three calcification outcomes were analyzed separately: sheet calcification area (%), nodular calcification area (%), and total calcification area (%). To reduce skewness, calcification outcomes were log2-transformed after adding a small constant (ε = 1×10⁻⁶). Regression coefficients were back-transformed and reported as fold-changes (2^β) with 95% confidence intervals.

### Fine-Gray regression models

Fine-Gray regression model accounting for competing risk was used for both cardiac death and major coronary events.^42^ The event indicator was defined as: 0 = alive/censored, 1 = cardiac death (/coronary event), 2 = other death. Cumulative incidence functions (CIFs) for cardiac death and major coronary events were estimated across calcification tertiles using the non-parametric method of Fine and Gray as implemented in the cmprsk R package, and differences between CIFs were assessed using Gray’s test. Associations between calcification tertiles and cardiac mortality were estimated with Fine-Gray subdistribution hazard regression (cmprsk::crr), reporting subdistribution hazard ratios (sHRs) with 95 % confidence intervals. Adjustments for age, sex, smoking status, diabetes, hypertension, kidney dysfunction, and low-density lipoprotein levels were included in the models. Prespecified sex-stratified analyses with similar covariate adjustments were additionally performed in men and women separately. In addition, Fine-Gray regression model with an additional interaction variable between calcification group and sex was used to test the statistical significance of the sex-dependency of the association observed.

## Results

### Study population

A total of 479 patients with available plaque samples were included and followed from 30 days after CEA. At baseline, median age was 71 years in women and 69 years in men (Table 1).

**Table 1.** Study population description and comparison of baseline characteristics between the sexes. The p-values are represented as * < 0.05, ** < 0.01, and *** < 0.001.

|  | Men (n = 325) | Women (n = 154) | p |  |
| --- | --- | --- | --- | --- |
| Age | 68.95 ± 8.19 | 71.15 ± 8.84 | p=0.003 | ** |
| BMI | 27.37 ± 3.91 | 28.03 ± 5.72 | p=0.066 |  |
| Waist measurement | 102.31 ± 10.48 | 97.25 ± 14.34 | p<0.001 | *** |
| Symptomatic plaque† | 209 (63.0 %) | 111 (70.7 %) | p=0.056 |  |
| Concomitant diseases |  |  |  |  |
| Kidney dysfunction | 53 (16.0 %) | 25 (15.9%) | p=0.552 |  |
| Coronary artery disease (CAD) | 144 (34.3%) | 37 (23.6%) | p<0.001 | *** |
| History of CABG | 51 (15.4%) | 9 (5.7 %) | p=0.001 | *** |
| History of myocardial infarction | 79 (23.8 %) | 14 (8.9%) | p<0.001 | *** |
| Hypertension | 271 (81.6%) | 127 (80.9%) | p=0.468 |  |
| Chronic heart failure | 32 (9.6%) | 18 (11.5%) | p=0.318 |  |
| Atrial fibrillation | 63 (19.0 %) | 20 (12.7%) | p=0.054 |  |
| Peripheral artery disease | 62 (18.7 %) | 23 (14.6%) | p=0.166 |  |
| Diabetes | 116 (34.3 %) | 49 (31.2%) | p=0.399 |  |
| Smoking status | 124 (37.3%) | 56 (35.7%) | p=0.518 |  |
| Medications |  |  |  |  |
| Low-dose aspirin | 171 (51.5%) | 80 (51.0%) | p=0.493 |  |
| Clopidogrel | 47 (14.2%) | 28 (17.8%) | p=0.179 |  |
| Dual antiplatelet therapy (ASA+CLO) | 12 (3.6%) | 10 (6.4%) | p=0.128 |  |
| Warfarin | 57 (17.2%) | 21 (13.4%) | p=0.175 |  |
| Lipid lowering agent | 327 (96.7 %) | 148 (94.3%) | p=0.107 |  |
| ACEi or ATRb | 248 (74.7%) | 109 (69.4%) | p=0.560 |  |
| Beta blocker | 181 (54.5%) | 90 (57.3%) | p=0.326 |  |
| Calcium channel blocker | 114 (34.3 %) | 62 (39.5%) | p=0.157 |  |
| Diabetes medication | 84 (25.3%) | 42 (26.8%) | p=0.369 |  |
| Lab measurements |  |  |  |  |
| Total cholesterol (mmol/L) | 5.65 ± 1.98 | 6.10 ± 1.23 | p=0.005 | ** |
| LDL cholesterol (mmol/L) | 3.65 ± 1.02 | 3.91 ± 1.12 | p=0.005 | ** |
| hs-CRP (mg/L) | 5.27 ± 13.00 | 5.44 ± 9.09 | p=0.434 |  |
| ApoB/ApoA | 0.59 ± 0.28 | 0.60 ± 0.27 | p=0.389 |  |
| P-Fibrinogen (g/L) | 3.94 ± 0.91 | 4.03 ± 0.93 | p=0.159 |  |

ASA+CLO, Low-dose aspirin and clopidogrel
ACEi, Angiotensin-converting enzyme inhibitor
ATRb, Angiotensin receptor blocker
hs-CRP, high-sensitivity C-reactive protein
† Patient had suffered ipsilateral ischemic stroke, retinal infarct, TIA or amaurosis fugax within 180 days.

At baseline, women were less likely to have suffered myocardial infarction (8.9% vs. 23.8%, p < 0.001) or to have undergone CABG (5.7% vs. 15.4%, p = 0.001) and had smaller waist circumference (97.3 cm vs. 102.3 cm, p < 0.001). In contrast, women had less favorable lipid profiles compared with men, including higher total cholesterol (6.1 mmol/L vs 5.7 mmol/L, p = 0.005) and LDL cholesterol levels (3.9 vs. 3.7, p = 0.005).

### Carotid plaques were more calcified in women than in men

Carotid plaques were more extensively calcified in women than in men. Median total plaque calcification area was higher in women (12.7% vs. 10.8%, p = 0.002), as were sheet calcification area (6.6% vs 5.3%, p < 0.001) and nodular calcification area (median 2.2% vs 1.9%, p = 0.030) (Table 2, and Figure S2-S3).

**Table 2.** Sex-stratified calcification quantification results. Results are presented as median and inter-quartile ranges.

|  | Men (n=325) | Women (n=154) |  |
| --- | --- | --- | --- |
| Nodular calcification area (%) | 1.9 (0.2-7.2) | 2.2 (0.4-7.2) | p = 0.030 |
| Sheet calcification area (%) | 5.3 (1.1-14.7) | 6.6 (1.5-16.2) | p < 0.001 |
| Total calcification area (%) | 10.8 ± (1.9-24.9) | 12.7 (2.9-26.6) | p = 0.002 |

### Plaque calcification was associated with baseline atherosclerotic disease

In age-adjusted linear regression analyses of the whole cohort, plaque total calcification was associated with baseline CAD (β = 0.73, 95% CI 0.07–1.39; p = 0.031). A similar association was observed for sheet calcification (β = 0.88, 95% CI 0.03–1.73; p = 0.042), whereas nodular calcification was not associated with CAD (β = 0.78, 95% CI −0.30–1.87; p = 0.16). These estimates correspond to approximately 1.7–1.8-fold higher plaque calcification proportions per presence of CAD.

In sex-stratified analyses, effect estimates of calcification for CAD were comparable in women but did not reach statistical significance (total calcification: β = 0.78, 95% CI −0.09–1.66; p = 0.077; sheet calcification: β = 1.35, 95% CI −0.01–2.71; p = 0.052), and no significant associations were observed in men (total calcification: β = 0.69, 95% CI −0.18–1.56; p = 0.12; sheet calcification: β = 0.70, 95% CI −0.38–1.77; p = 0.20).

Carotid plaque nodular calcification was associated with baseline PAD in the whole cohort (β = 1.58, 95% CI 0.29–2.89, p = 0.017). Patients with PAD had an approximately threefold higher proportion of nodular calcification in carotid plaques compared with patients without PAD. Sex-stratified analyses were limited by the small number of PAD cases at baseline and did not reach statistical significance (women, N=23: β = 1.76, 95% CI −0.47 to 3.99, p = 0.12; men, N=62: β = 1.51, 95% CI −0.09 to 3.12, p = 0.065). Associations between PAD and sheet or total calcification in the overall cohort were not statistically significant (total calcification: β = 0.72, 95% CI −0.07 to 1.53, p = 0.078; sheet calcification: β = 0.85, 95% CI −0.18 to 1.88, p = 0.11).

### Plaque calcification was associated with baseline cardiac morbidity

In the overall cohort, plaque calcification was associated with baseline chronic heart failure (CHF), irrespective of calcification subtype. Total calcification was associated with CHF (β = 1.32, 95% CI 0.31–2.32; p = 0.010), corresponding to approximately 2.5-fold higher proportion of calcification. Similar associations were observed for sheet calcification (β = 1.44, 95% CI 0.10–2.73; p = 0.028) and nodular calcification (β = 1.85, 95% CI 0.23–3.48; p = 0.026).

In sex-stratified analyses, total calcification remained associated with CHF in both women (β = 1.12, 95% CI 0.00–2.23; p = 0.049) and men (β = 1.43, 95% CI 0.01–2.85; p = 0.047), whereas associations for sheet and nodular calcification separately were attenuated (men: sheet β = 1.58, 95% CI 0.89–10.01, p = 0.144; nodular β = 1.60, p = 0.077; women: sheet β = 1.21, p = 0.173; nodular β = 2.29, p = 0.065).

Total calcification (β = 0.93, 95% CI 0.10–1.77; p = 0.028) and sheet calcification (β = 1.29, 95% CI 0.22–2.36; p = 0.018) were associated with atrial fibrillation in the overall cohort, whereas nodular calcification was not (β = 0.61, 95% CI −0.74–1.96; p = 0.377). Statistical significance of these associations did not retain in sex-stratified analyses (Table S1).

Nodular calcification was associated with baseline hypertension in men (β = 1.76, 95% CI 0.13–3.38, p = 0.035), whereas sheet calcification was associated with hypertension in women (β = 1.59, 95% CI 0.16–3.02, p = 0.029). Total calcification was not associated with hypertension in the overall cohort or in either sex (Table S1).

### Number of endpoint events during follow-up

During a median follow-up of 10.7-years, 253 patients (49.1 %) died, of whom 70 deaths (27.7 %) were cardiac. Among cardiac deaths, 25 (35.7 %) occurred in women. Of the cardiac deaths, 20 (28.6 %) were further categorized as sudden cardiac death and 9 (12.9 %) as death due to acute myocardial infarction. A total of 76 patients (15.9 %) experienced a major coronary event, and 69 (14.4 %) experienced a MALE event. Event numbers stratified by calcification tertiles are presented in Tables S2-S4.

### Nodular calcification was associated with increased risk of cardiac death

Cumulative incidence of cardiac death differed significantly between patients in the middle (T2) and high (T3) tertiles compared with the lowest tertile (T1) for plaque nodular calcification (Gray’s test p = 0.001; Figure 2). The association appeared threshold-type, with no significant difference between T2 and T3 (sHR 0.89, 95% CI 0.53─1.51, p = 0.670).

**Figure 1.**
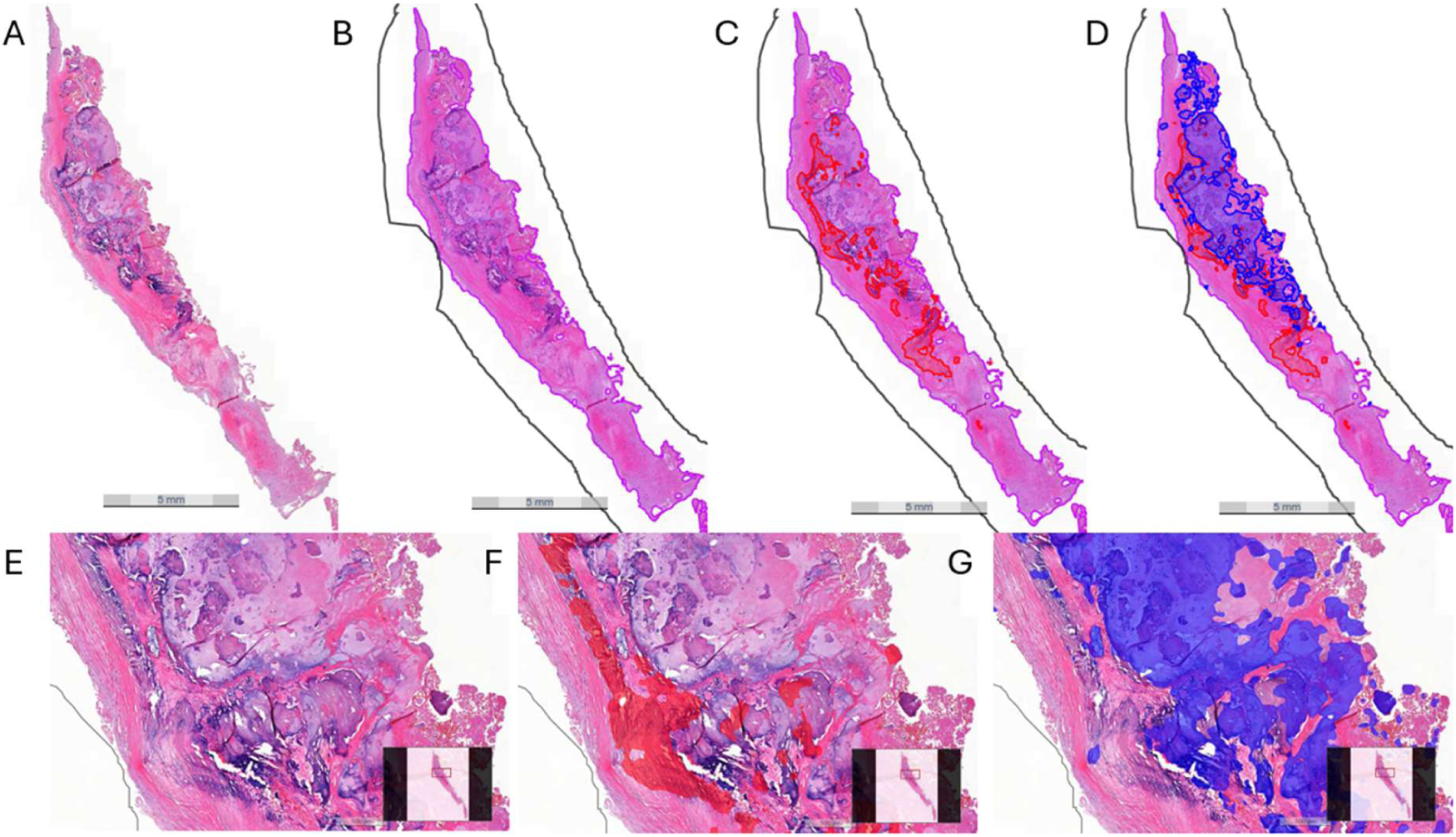
Representative example of AI-based calcification quantification in (A and E) HE-stained longitudinal section of carotid plaque. (B) The total tissue area was quantified in mm² (purple). The algorithm identified and quantified areas of sheet calcification (red, C, and G) and nodular calcification (blue, D, and F). (D) Total plaque calcification area (%) was calculated as the sum of sheet and nodular calcification areas (D, %; red + blue) as proportion of the tissue area (%; A).

**Figure 2.**
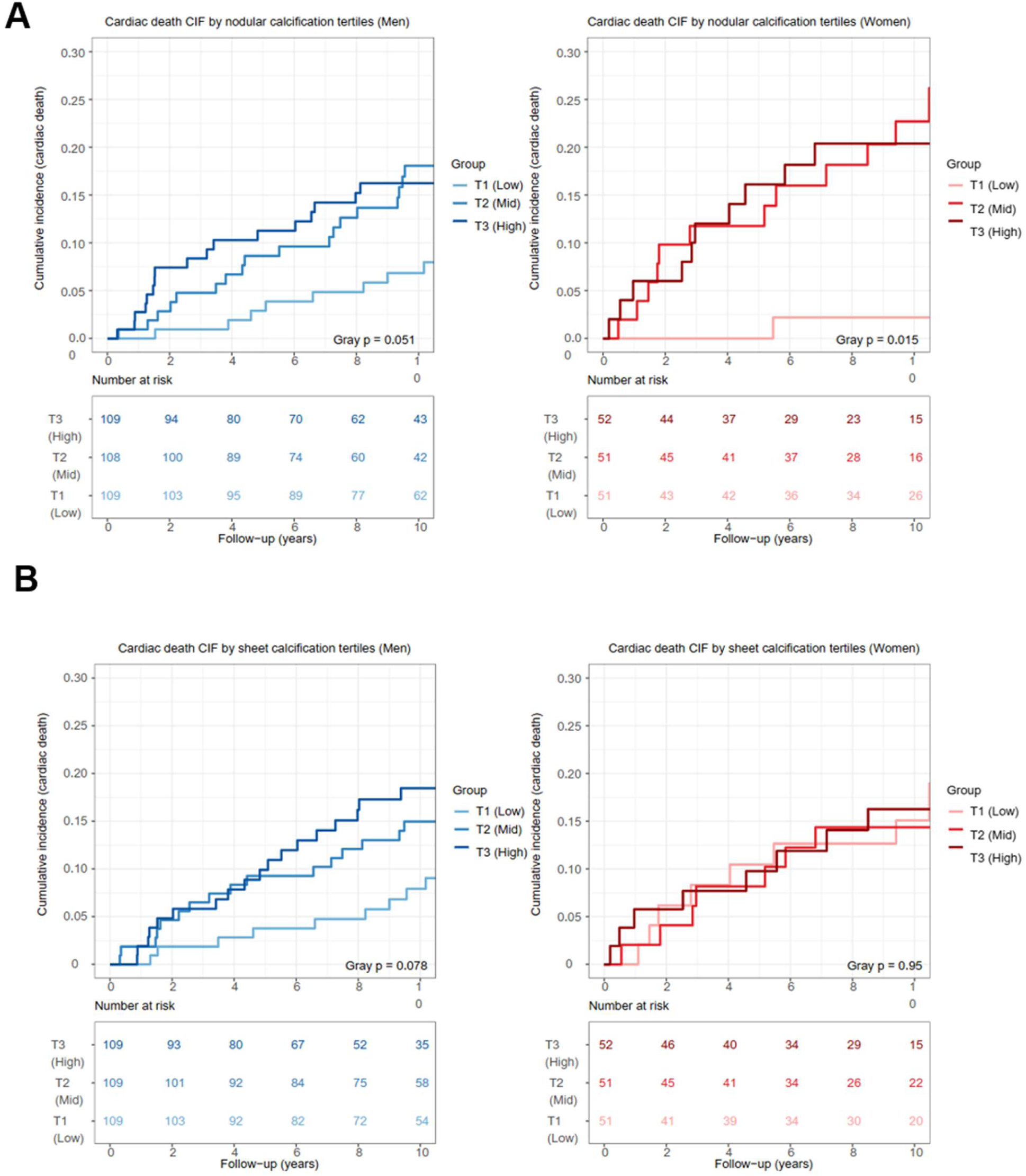
Sex-stratified cumulative incidence function (CIF) curves for cardiac death during 10-year follow-up according to carotid plaque (A) nodular calcification burden and (B) sheet calcification burden. Low (T1), medium (T2), and high (T3) calcification groups are shown separately for women and men. Numbers at risk are presented below the curves.

Patients in T2 and T3 had a 2.9-fold and 2.6-fold higher risk of cardiac death, respectively, compared with T1 in Fine-Gray regression models accounting for competing risks and adjusted for age, sex, BMI, LDL cholesterol, hypertension, kidney dysfunction, diabetes, and smoking status (sHR 2.87, 95% CI 1.39─5.91, p = 0.004 for T2; sHR 2.55, 95 % CI 1.24─5.27, p = 0.011 for T3). Cumulative incidence rates of cardiac death were 5.4% in T1, 19.6% in T2 and 17.6% in T3.

In sex-stratified analyses, the association between nodular calcification and cardiac death was more pronounced in women. The risk of cardiac death was substantially increased in T2 and T3 compared with T1 (Gray’s test p = 0.015; sHR 5.63, 95% 1.31─24.17, p = 0.020 for T2; sHR 4.93, 95 % CI 1.14─21.41, p = 0.033 for T3).

In men, the association was attenuated but was near statistical significance in covariate adjusted Fine-Gray model (Gray’s test p = 0.051; sHR 2.05, 95% 0.85─4.92, p = 0.11 for T2, and sHR 2.30, 95 % CI 0.98─5.40, p = 0.056 for T3). Sex-specific cumulative incidence rates were 3.9%, 23.5%, and 21.2% for women and 7.4%, 16.7%, and 17.4 % for men across nodular calcification tertiles T1-T3, respectively.

Among covariates, baseline age associated with cardiac death in both women (sHR 1.05, 95 % CI 1.01─1.10, p = 0.024, Table S6) and men (sHR 1.05, 95 % CI 1.01─1.10, p = 0.022, Table S7). In men, cardiac death was additionally associated with baseline hypertension diagnosis (sHR 0.43, 95 % CI 0.21─0.90, p = 0.025; Table S7), diabetes (sHR 2.11, 95 % CI 1.07-4.17, p = 0.032), chronic kidney dysfunction (sHR 2.57, 95 % CI 1.39─4.77, p = 0.003).

### The highest tertile of sheet calcification was associated with cardiac death in men

In women, sheet calcification was not associated with cardiac death (Table S12). In contrast, men in the highest tertile (T3) had an increased risk of cardiac death compared with those in the lowest tertile (T1) in Fine-Gray regression models accounting for competing risks and adjusted for age, BMI, LDL cholesterol, hypertension, diabetes, and smoking (sHR 2.3, 95 % CI 1.0─5.1, p = 0.045; Figure 2). No significant difference in cumulative incidence was observed between T2 and T1 (Table S13).

### Total calcification did not predict cardiac mortality

Total calcification was not associated with cardiac mortality in either the overall cohort or in sex-stratified analyses.

### Total calcification was associated with major coronary events in women

During follow-up, four women (7.8%) both in T1 and T2 and 12 women (23.5%) in T3 experienced a major coronary event (Gray’s test p = 0.034; Tables S2–S4; Figure 3).

**Figure 3.**
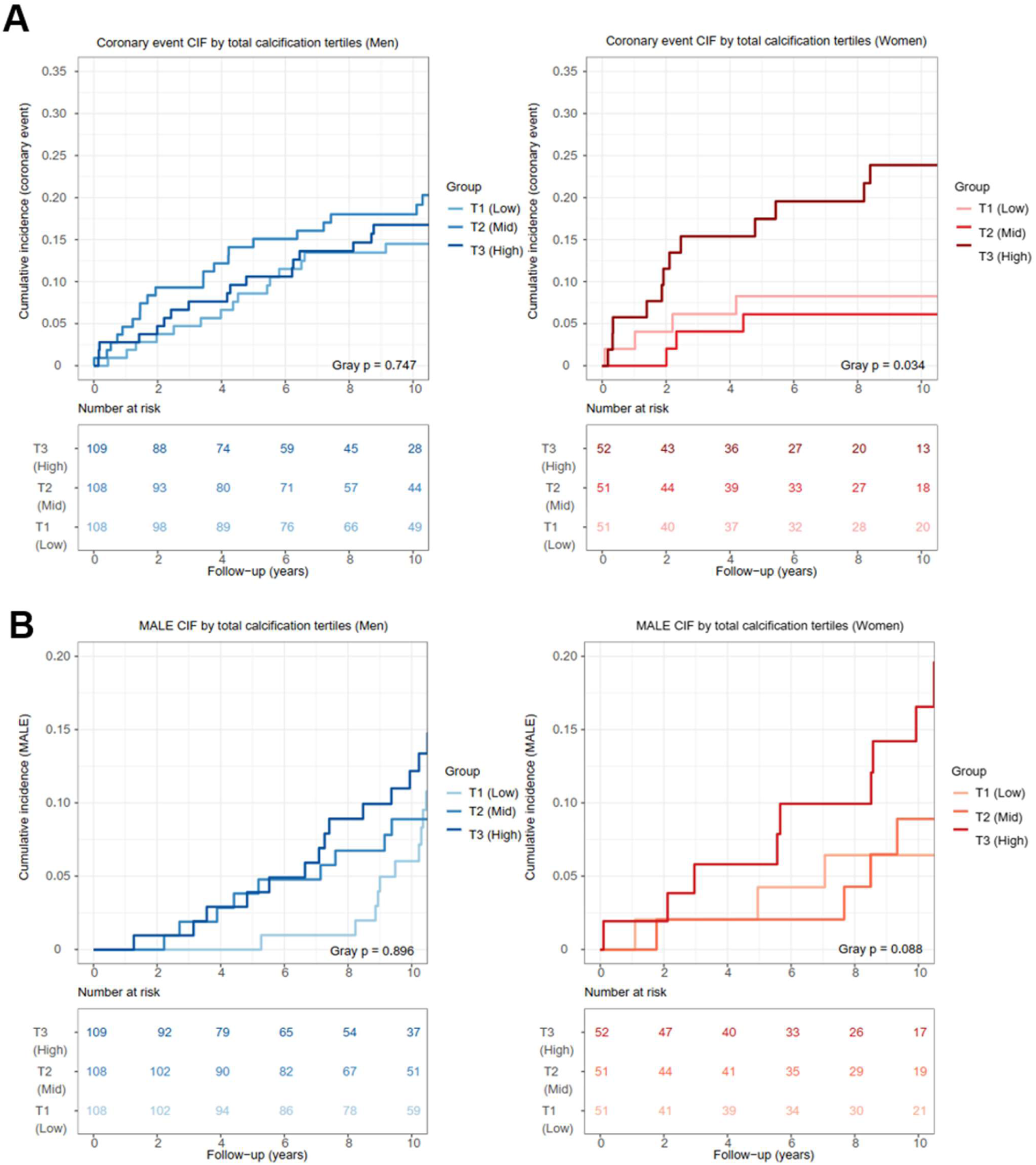
Sex-stratified cumulative incidence function (CIF) curves for (A) major coronary events and (B) major adverse limb events (MALE) during 10-year follow-up according to carotid plaque total calcification burden. Low (T1), medium (T2) and high (T3) calcification groups are shown separately for women and men. Numbers at risk are presented below the curves.

In multivariable Fine–Gray regression models, no difference was observed between T1 and T2 (sHR 0.95, 95% CI 0.25–3.56, p = 0.94), whereas women in T3 showed a trend toward increased risk of major coronary event (sHR 3.09, 95% CI 0.91–10.52, p = 0.071). In an exploratory analysis combining T1 and T2, T3 was associated with a higher risk of major coronary events (sHR 3.17, 95% CI 1.19–8.49, p = 0.021).

In men, no association was observed (sHR 1.39, 95% CI 0.73–2.64, p = 0.31 for T2; HR 1.26, 95 % CI 0.64─2.48, p = 0.51 for T3). A sex interaction term supported sex-specific differences in this association (p = 0.033). Neither nodular nor sheet calcification alone was associated with major coronary events.

Among covariates, baseline age was inversely associated with major coronary events in men (sHR 0.95, 95 % CI 0.92–0.98, p = 0.001; Table S9), but not in women (sHR 0.99, 95 % CI 0.95–1.04, p = 0.81; Table S8). Baseline CAD was associated with major coronary event in women (sHR 3.12, 95 % CI 1.13–8.60, p = 0.027; Table S8), but not in men (sHR 1.46, 95 % CI 0.78–2.74, p = 0.23; Table S9).

### Total calcification was associated with MALE in women

During follow-up, MALE occurred in 7.8% of women in T1–T2 and 21.2% in T3 (Gray’s test p = 0.028). In women, total calcification was associated with MALE at borderline statistical significance (T3, sHR 3.27, 95% CI 1.0–10.76, p = 0.051).

When low and middle total calcification tertiles (T1 and T2) were combined, T3 was associated with increased risk of MALE (sHR 2.80, 95% CI 1.12–6.95, p = 0.027; Figure 3). For calcification subtypes, moderate nodular calcification (T2) was associated with increased MALE risk compared with the lowest tertile in female patients (sHR 3.96, 95% CI 1.01–15.46, p = 0.048; Gray’s test p = 0.053). No association was observed for sheet calcification.

In men, plaque calcification was not associated with MALE (Tables S16-S18). Among covariates, diabetes was associated with MALE in women (sHR 3.27, 95 % CI 1.17–9.14, p = 0.023, Table S14) but not in men (sHR 1.38, 95 % CI 0.76–2.52, p = 0.29; Table S16), whereas, in men only, chronic kidney disease and smoking were associated with MALE event (sHR 2.59, 95 % CI 1.30–5.16, p = 0.007 for chronic kidney disease; sHR 0.54, 95 % CI 0.29–1.00, p = 0.05 for smoking status; Table S16). Baseline age was inversely associated with subsequent MALE event in men only (sHR 0.96, 95 % CI 0.93–1.00, p = 0.032; Table S16).

## Discussion

In this study, carotid plaque calcification and its morphological subtypes were differentially associated with systemic cardiovascular disease and long-term outcomes after carotid endarterectomy. The principal finding was that nodular calcification was strongly and independently associated with long-term cardiac mortality, with larger effect estimates in women (). In contrast, total calcification was associated with cardiovascular comorbidity, and in women, with subsequent coronary and limb events, but did not predict cardiac mortality. These findings support the concept that vascular calcification is a heterogenous plaque feature and that its clinical meaning depends on morphological phenotype and may differ by sex.

The presence of carotid plaque has previously been consistently associated with incident CAD.^43,44^ A meta-analysis further demonstrated a correlation between carotid calcification and severity of CAD.^45^ However, studies evaluating carotid calcification as a marker of subsequent cardiovascular events have yielded conflicting results, with some reporting no independent association between carotid calcification and coronary events,^46,47^ and others describing associations with cardiac death, myocardial infarction, or systemic vascular events.^48–50^ Our findings extend this literature by showing that histologically defined calcification morphology may partly explain these discrepancies. In particular, nodular calcification, rather than total calcification, identified patients at increased risk of cardiac death, whereas total calcification was more closely related to coronary and peripheral event risk in women. Thus, calcification burden alone may be insufficient for risk assessment, and evaluation of calcification subtype may provide additional prognostic information.

Our study also expands current knowledge on the relationships between carotid plaque calcification and cardiac morbidity. Previous studies have linked carotid calcification with hypertension^51^ and incident atrial fibrillation.^52^ Consistent with these observations, we found associations between calcification measures and both hypertension and atrial fibrillation. In addition, total calcification was associated with chronic heart failure in both women and men. To our knowledge, this association between carotid plaque calcification and heart failure has not previously been reported. Although this finding requires confirmation, it supports the concept that calcification in carotid plaques may reflect systemic cardiovascular disease beyond the local cerebrovascular territory.

A central observation was that nodular calcification predicted cardiac death over long-term follow-up. Importantly, this association was not paralleled by a corresponding increase in major coronary events, and most cardiac deaths were attributable to chronic cardiac conditions rather than acute myocardial infarction. Together, these findings suggest that nodular calcification in carotid artery may reflect advanced systemic cardiovascular disease and chronic myocardial disease severity rather than coronary plaque instability. This interpretation is further supported by the observed associations between nodular calcification and baseline cardiac comorbidities, including chronic heart failure and atrial fibrillation.

In contrast, total calcification proportion of carotid plaque tissue area was not associated with cardiac mortality, underscoring that aggregate calcification does not fully capture prognostic risk. Rather, the divergent associations of nodular and total calcification suggest that calcification morphology and calcification burden reflect partly distinct dimensions of systemic atherosclerotic disease. Nodular calcification may mark disease severity and adverse long-term prognosis, whereas total calcification appears to reflect disease extent and susceptibility to coronary and peripheral events, particularly in women.

Our results also provide insight into the relationship between carotid plaque calcification and systemic atherosclerosis involving coronary and peripheral vascular beds. Total calcification was associated with baseline CAD, and nodular calcification with baseline PAD, suggesting that carotid plaque calcification may reflect diffuse atherosclerotic involvement across multiple vascular territories. During follow-up, exploratory sex-stratified analyses suggested that high total calcification was associated with major coronary events and major adverse limb events in women, whereas corresponding associations were not observed in men. In contrast, nodular calcification predicted cardiac death in the whole cohort, with larger effect estimates in women, while high sheet calcification was associated with cardiac death in men.

The observed sex-related heterogeneity should be interpreted cautiously but is biologically plausible. Women typically develop atherosclerotic disease later than men, but disease burden and morbidity increase rapidly after menopause.^27,28^ Previous studies have also reported sex-related differences in carotid and coronary plaque characteristics.^31–36^ The background mechanisms behind these sex-related differences remain elusive. Estrogen might play a role in addition to sex-related differences in genetics and immunological factors.^43^ In our cohort, women had a lower baseline prevalence of diagnosed CAD despite less favorable lipid profiles, raising the possibility that carotid plaque characteristics may help identify subclinical or underdiagnosed cardiovascular disease in this population. Overall, these findings indicate that carotid plaque calcification burden differs between sexes and that both its clinical correlates and prognostic implications are sex related, emphasizing the importance of reporting sex-stratified analyses and investigating sex-specific mechanisms in future studies.

From a clinical perspective, these findings suggest that histological characterization of carotid plaque calcification could provide prognostic information beyond traditional cardiovascular risk factors. Patients with high nodular calcification burden may represent a subgroup at increased risk of cardiac death related to chronic cardiac disease. Current clinical practice does not incorporate plaque morphology into cardiovascular risk assessment after carotid endarterectomy, ^9,53^ but our results support further investigation into whether calcification phenotype could improve risk stratification and guide targeted cardiovascular evaluation.

The strengths of this study include a well-characterized prospective cohort, long-term follow-up, and quantitative AI-based histological assessment of calcification subtypes. However, several limitations should be acknowledged. The number of outcome events, particularly in sex-stratified analyses, was limited, which may reduce statistical power and increase uncertainty around effect estimates. Calcification assessment was based on a single plaque specimen. In addition, the observational design precludes causal inference, and residual confounding cannot be excluded.

In conclusion, carotid plaque calcification is differentially associated with cardiovascular outcomes depending on its morphological subtype. Nodular calcification is a strong predictor of long-term cardiac mortality and may reflect advanced systemic cardiovascular disease, whereas total calcification burden is associated with coronary and peripheral arterial events, particularly in women. These findings emphasize the importance of plaque characterization and suggest that calcification phenotypes may provide clinically relevant insight into cardiovascular risk beyond conventional risk factors.

## Data Availability

The data underlying this study contain sensitive patient-level health information and are not publicly available due to ethical, legal, and data protection restrictions. Data access may be considered upon reasonable request to the corresponding author, subject to approval by the relevant institutional authorities and ethics committee, and in accordance with applicable data protection regulations.

## Non-standard Abbreviations and Acronyms

ACEi: angiotensin-converting enzyme inhibitor
AI: artificial intelligence
ASA: low-dose aspirin / acetylsalicylic acid
ATRb: angiotensin receptor blocker
BMI: body mass index
CABG: coronary artery bypass grafting
CAD: coronary artery disease
CEA: carotid endarterectomy
CHF: chronic heart failure
CIF: cumulative incidence function
CLO: clopidogrel
HE: hematoxylin–eosin
HeCES2: Helsinki Carotid Endarterectomy Study 2
hs-CRP: high-sensitivity C-reactive protein
LDL: low-density lipoprotein
MALE: major adverse limb event
PAD: peripheral artery disease
PCI: percutaneous coronary angioplasty
SD: standard deviation
sHR: subdistribution hazard ratio
T1: lowest/low tertile
T2: middle/medium tertile
T3: highest/high tertile

